# Is the messenger more important than the message? A theory of change for the IFRC Network’s risk communication and community engagement intervention for vaccination in Nigeria and Ethiopia

**DOI:** 10.64898/2026.04.29.26352040

**Authors:** Rose Burns, Yashua Alkali Hamza, Aminu M. Dukku, Yohannes Mulugeta, Ombretta Baggio, Gefra Fulane, Monica Posada, Lilian Adeogba, Abraham Tilahun, Alex Odlum, Karl Blanchet, Luisa Enria

**Affiliations:** London School of Hygiene & Tropical Medicine, United Kingdom; Childcare and Wellness Clinics Abuja, Nigeria; Bayero University, Kano, Nigeria; Addis Ababa University, Addis Ababa, Ethiopia; International Federation of Red Cross and Red Crescent Societies, Geneva, Switzerland; International Federation of Red Cross and Red Crescent Societies Nigeria, Abuja, Nigeria; International Federation of Red Cross and Red Crescent Societies Ethiopia, Addis Ababa, Ethiopia; University of Geneva, Geneva, Switzerland

**Keywords:** community engagement, risk communication, community engagement, vaccination, Nigeria, Ethiopia, theory of change

## Abstract

**Introduction:** Community engagement is increasingly seen as essential within vaccination programming to improve uptake of vaccines, build trust and foster community ownership. Yet the goals and mechanisms of such interventions are often contested or not explicit. This article aims to address this by examining how those directly involved in implementing community engagement understand its intended aims and outcomes. We use as a case study a risk communication and community engagement (RCCE) intervention implemented by the Nigeria and Ethiopia Red Cross/Red Crescent with support from the IFRC for COVID-19 vaccination.

**Methods:** We conducted 41 interviews, 12 participatory workshops and citizen ethnography in Dire Dawa, Ethiopia and Kano, Nigeria including with Red Cross/Red Crescent and vaccination staff, vaccine users and community members. We explored how participants understood the RCCE intervention’s ‘theory of change’, including how it was *expected* to work, for whom, under what circumstances, and why.

**Results:** Participants described RCCE activities as a mix of ‘two-way’ (such as house-to-house visits) and ‘mass’ approaches (such as media campaigns). These interventions were primarily seen as enhancing vaccine knowledge and countering misinformation. Key ‘mechanisms’ included vaccine users’ willingness to act on the information provided, however this was heavily influenced by the credibility and trustworthiness of the bearers of vaccine information. While feedback mechanisms existed, communities were not involved in designing vaccination strategies. Efforts were shaped by a context with unpredictable vaccination campaigns, supply constraints and parallel RCCE efforts by community actors.

**Conclusions:** We show that in this theory of change ‘messengers’ are more influential than the messages themselves. By developing a theory of change with our participants, we highlight the lack of clarity within the sector regarding the definition and expected impact of community engagement and reveal a gap between community engagement practice on the ground and normative goals such as co-production and dialogue.

**Key messages:** *What is already known on this topic:* Risk communication and community engagement (RCCE) is widely used to improve vaccination programmes, but there is limited evidence on how such interventions are expected to work, for whom, under what circumstances and why.

*What this study adds:* We examined how those involved in implementing a Red Cross/Red Crescent and IFRC RCCE intervention in Ethiopia and Nigeria understood the aims and outcomes of this intervention. Whilst our study participants believed their main goal was to share correct vaccine information to counter rumours, we found that trust in the people who delivered the information was often more important than the information delivery itself.

*How this study might affect research, practice or policy:* Our research highlights tension between normative ideals of community engagement and how it is understood and practiced by implementors in humanitarian contexts, and underscores the need to move beyond knowledge-deficit approaches.

## Introduction

Implementing vaccination programmes in humanitarian contexts is complex, involving logistical constraints, limited health system capacity and vaccine hesitancy.

Community engagement is increasingly seen as a core activity in humanitarian interventions, and essential for improving vaccine uptake in complex emergencies ^1^. Yet, it’s definition and operationalisation remains contested and it encompasses a wide spectrum of activities and goals ^2^. It is broadly understood as efforts to bring together “traditional, community, civil society, government, and opinion groups and leaders; and expanding collective or group roles in addressing the issues that affect their lives”^3^. Community engagement may be framed either as an ‘end in itself’—for example, to empower and strengthen communities so they can address issues that affect them—or as a ‘means to an end’ aimed at achieving outcomes in areas such as health, education, or vaccination coverage^4^.

In the humanitarian sector, community engagement includes a range of (overlapping) typologies. ‘Risk communication and community engagement’ (RCCE) focuses on engaging, informing and sharing key information with communities often during crises such as outbreaks. ‘Community engagement and accountability’ focuses on promoting accountability to affected populations through dialogue on the appropriateness and effectiveness of interventions as well as engaging people in intervention planning ^5,6^. The *Minimum Quality Standard* describe six core standards for community engagement: ^4^: i. ‘participation’, meaning communities assess their own needs and participate in developing humanitarian initiatives; ii. ‘empowerment and ownership’ which are defined as both processes and an outcomes; iii. ‘inclusion’ of underrepresented or marginalised groups; iv. Two-way communication to give and receive accurate information; v. ‘adaptability and localisation’ meaning changes are made based on local settings’; and vi. ‘building on local capacity’ meaning the building on the existing skills and resources of communities and the local groups that serve them.

Alongside debates about the forms that community engagement can take, there is also the question of it’s intended outcome. Some literature links community engagement interventions to outcomes such as improved vaccine acceptance and coverage rates. Approaches like working with trusted actors, such as in school-based vaccination programs, or engaging community volunteers or community health workers can increase vaccine uptake, especially among hard-to-reach populations ^7,8^. Engaging traditional and religious leaders who wield influence to address misinformation, and using tailored communication strategies have also been effective in addressing vaccine hesitancy ^9–11^. Beyond these instrumental goals, some argue that community engagement is also associated with more emancipatory goals, such as shifting power to communities, generating accountability, co-production for humanitarian programming, or governance more broadly ^12^.

### Existing evidence on how community engagement works and its impact in humanitarian contexts

The mechanisms through which community engagement activities lead to outcomes (and what those outcomes should be) are often assumed but not made explicit by humanitarian actors. Evidence gaps remain on how and why specific interventions are effective. A realist scoping review identified several promising mechanisms for community engagement interventions in a crisis response ^13^, including community structured dialogue; partnership coordination and the use of existing structures, for example, known community and opinion leaders and established health facilities; as well as empowering local people from the affected community to be involved in the response. It also highlighted the value of citizen science, where communities co-design, implement, and produce data, generating useful insights and better addressing local needs. Community engagement interventions have aimed to achieve various outcomes including the development of sustainable structures, building trust, development of social resilience, community development, programme development, and ultimately the improvement of health indicators ^13–17^.

### Theory of change

Against this backdrop, our research interrogates the mechanisms and intended aims of community engagement by drawing on the experiences of implementors and intended beneficiaries. We used a ‘theory of change’ to make explicit how Red Cross/Red Crescent (RC) staff and participating communities/actors perceived the CE/RCCE intervention to work, for whom and under what circumstances. This aimed to surface underlying theories about engagement and make them explicit, define current practice, and highlight gaps between policy and how this is understood and practiced on the ground.

A realist approach to a ‘theory of change’ explains how programme activities are understood to generate as series of results (‘outcomes’) through ‘mechanisms’ within a specific context (‘context’) ^18,19^. ‘Mechanisms’ refer to the participants’ responses to the programme activities and link activities to outcomes. The focus is not on whether a programme achieved outcomes or not, but rather on how those implementing the programme expected it to achieve outcomes.

In this study, we describe how the intervention was planned, delivered, and taken up by communities as well as the mechanisms through which the intervention was expected to improve vaccination uptake, and the contextual factors affecting implementation and outcomes in Ethiopia and Nigeria. This paper reflects staff understanding of the programme theory, rather than an ‘ideal’ for how community engagement should operate. It highlights assumptions from frontline workers and communities, and uses these to ask normative questions about what community engagement *should* be. Our objectives were to understand, from the perspective of our participants: i. how is the IFRC Network’s RCCE intervention perceived to work ii. what are the IFRC Network’s community engagement approaches, their expected impacts, and the ways these impacts are intended to be achieved iii. what are the barriers and facilitators to implementing RCCE activities.

## Methods

### Study setting

This study was conducted in Kano, Nigeria and Dire Dawa, Ethiopia. Kano, Nigeria’s second largest city, is in the predominantly Muslim North. A diphtheria outbreak that began in 2022 was ongoing during our research in 2023-2024, with Kano among the most affected states ^20^. Routine childhood vaccination coverage is low, with mean coverage for BCG, Penta 1 and Penta 3 just over 50%^21,22^. Dire Dawa, in Eastern Ethiopia has comparatively higher routine coverage at 66% ^23^. As of December 2023, 38% of Ethiopia had completed a primary COVID-19 vaccine series, and 46% had received at least one dose ^24^. Ethiopia was also facing a large cholera outbreak at the time of our research.

Both sites face challenges achieving high vaccination coverage due to range of supply and demand factors including health care access, vaccine deployment infrastructure, political instability, and vaccine hesitancy and concerns ^25^. Both cities host substantial RC RCCE activities, including the Saving Lives and Livelihoods programme.

### Study partners

This study was conducted by a consortium of researchers, led by the Geneva Centre of Humanitarian Studies in collaboration with London School of Hygiene and Tropical Medicine, Addis Ababa University School of Public Health (Ethiopia), and Childcare and Wellness Clinics (Nigeria). The study was implemented in collaboration with International Federation of the Red Cross and Red Crescent Societies (IFRC) and the National Societies of Ethiopia and Nigeria.

### IFRC Risk Communication and Community Engagement: Saving Lives and Livelihoods programme

The IFRC is the world’s largest humanitarian network, made up of National Red Cross and Red Crescent Societies in each country, with the IFRC serving as the coordinating body. It is a global leader in community engagement and accountability. It’s network-wide commitments emphasise community participation, contextual understanding, handing over control of programmes to communities where possible; transparent communication; and establishing trusted feedback mechanisms^26^.

We focused on IFRC Network’s RCCE activities using the Saving Lives and Livelihoods (SLL) intervention as a case study ^27^. This $1.5 billion USD partnership between Africa Centre for Disease Control and Mastercard Foundation was launched in 2021 and aimed to purchase and deploy COVID-19 vaccines for at least 64 million people in Africa, targeting 70% coverage across the continent. IFRC and the National Red Cross and Red Crescent Societies are the lead partner for the RCCE pillar of the programme, which was implemented in both Kano, Nigeria, and Dire Dawa, Ethiopia, from 2021 to 2023. The programme reflects widely used RCCE approaches across the continent.

#### The RCCE pillar of SLL had the following objectives

1. To understand drivers and scale of vaccine hesitancy and develop a strategy to combat it
2. To implement RCCE strategies in selected countries
3. To set up systems and tools for effective communication within Member States

Key activities reported under the SLL programme were broadly similar in Kano and Dire Dawa sites, reflecting standard RCCE approaches used by the RC in many settings for vaccination. These are grouped under the IFRC Community Engagement and Accountability (CEA) intervention typologies:

**Table 1:**

| Risk communication and community engagement activities, Saving Lives and Livelihoods intervention<sup>1</sup> Kano and Dire Dawa |  |
| --- | --- |
| **‘Mass communication’** |  |
| i. | <u>Targeted communication/public entertainment events</u> including market ‘sensitisations’, road shows, DJs and recorded messages, town criers/announcers, and university campus sensitization. |
| ii. | <u>Broadcast media</u> campaigns using influential ambassadors. |
| **‘Two-way communication’** |  |
| iii. | Door-to-door visits by volunteers discussing COVID-19, routine immunisation, and HPV vaccines. Additional activities included identifying sick children and zero dose children. Discussions between volunteers and household members usually lasted several minutes , with 45-60 households reached per volunteer per day in a high-density area. |
| iv. | <u>Collection of community feedback:</u> questions and concerns/rumours about vaccines were collected using online/offline platform (replied to in real time). |
| **‘Community participation’** |  |
| v. | <u>Leadership consultation and community group meetings</u> with opinion leaders, led by stakeholder volunteers selected based on authority (e.g. religious leaders, women’s and youth groups, Kebele representatives, and industry park workers). |

### Understanding impact at IFRC

The IFRC has developed an impact framework for its community engagement activities to track and evaluate the (intended or unintended) changes resulting from this work (Figure 1). This enables teams to assess the effectiveness of CEA interventions, make data-informed decisions, and demonstrate accountability. Our study complements IFRC’s work on impact and aims to explain the link between activities and outputs (shown with the red arrows in Fig 1), specifically for the activities (circled in red) on the left of the diagram (mass communication, two-way communication and community participation). More broadly we sought to examine how such frameworks are translated in practice and understood by on the ground staff and communities implementing them.

**Figure 1:**
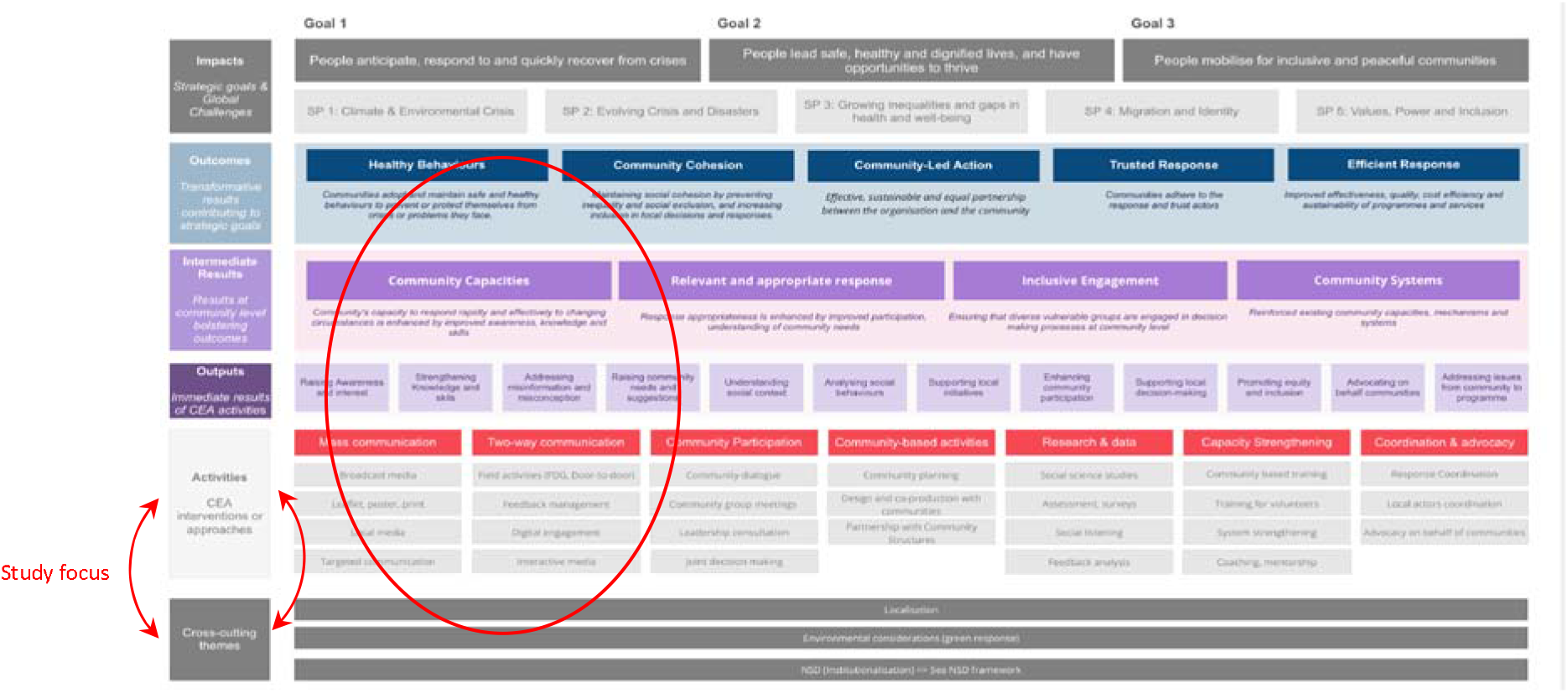
Impact framework at IFRC ^28^

### Sampling and recruitment

We used interviews, participatory workshops and observations to develop a theory of change. Interview participants included community leaders (e.g. religious leader, village leaders, influential community members), vaccine users, caretakers of vaccine users, health workers responsible for vaccination, government health officials, frontline vaccination staff (e.g. health extension workers), and RC/IFRC staff involved in RCCE activities across both countries.

Power mapping workshops were conducted separately with both community members and leaders. Theory of change workshops were held with IFRC and RC staff at study sites and in capital cities. Causal loop workshops were held with IFRC/RC staff, formal and informal health care workers and community stakeholders and vaccine users. IFRC and RC staff directly involved in RCCE programming were selected.

Participants were approached through IFRC Network, Addis Ababa University (AAU) and Childcare and Wellness Clinics (CWC) networks and invited to participate. Interviews and group activities were conducted in a private community space convenient for participants to reach, or in the case of some key informants, their place of work. Observation sites were selected in consultation with the IFRC Network to reflect a range of vaccination and community contexts.

### Data collection, management and analysis

Between January and May 2024, we conducted 41 interviews and 12 participatory workshops across both sites. This was complemented by several days of observations conducted by trained citizen ethnographers (10 in Dire Dawa and 12 in Kano) in vaccination sites and community locations. In Kano, ‘live’ RCCE activities conducted by the IFRC Network during a government led Diphtheria vaccine campaign.

**Table 2:** Number and characteristics of workshop participants.

| Activity | Number of workshops in Nigeria | Number of workshops in Ethiopia | Average number of participants per workshop | Total number of participants |
| --- | --- | --- | --- | --- |
| Power Mapping | 2 | 2 | 10 | 40 |
| Theory of Change | 2 | 2 | 10 | 40 |
| Causal Loop | 1 | 1 | 10 | 20 |
| TOTAL | 5 | 5 | 10 | 100 |

**Table 3:** Number and characteristics of interview participants.

| Interviewee Type | Nigeria | Ethiopia | Total |
| --- | --- | --- | --- |
| IFRC Network staff | 5 | 8 | 12 |
| Community leaders | 4 | 4 | 8 |
| Health officials and workers involved in vaccination | 5 | 8 | 13 |
| Community member recipients (or parent/ guardians of recipients) | 4 | 4 | 8 |
| TOTAL | 18 | 24 | 41 |

Interviews in Dire Dawa were conducted in Amharic by Ethiopian research assistants (RA), while workshops were co-facilitated by two RAs and two European researchers. In Kano, five Nigerian researchers conducted interviews and workshops in Hausa and English. Interviews averaged 45 minutes and were audio recorded, and workshops lasted 3 - 4 hours and were audio recorded in the case of Ethiopia and in the case of Nigeria detailed notes were taken. Participants were reimbursed for transport costs.

Theory of change topic guides generated visual diagrams following questions on the programme outcomes; preconditions, required activities and resources, and underlying assumptions, rationale and contextual conditions. Questions were adapted from De Silva et al ^29^.

Additional workshops developed causal loop diagrams (CLDs) to examine programme implementation and received in practice. Topic guides explored facilitators, barriers and potential leverage points for community engagement. Topic guides for power mapping workshops focussed on identifying and visually mapped key actors holding power and trust in participants’ communities, including specific individuals or groups(e.g. religious leaders/ nurses), producing details visual representations of influence within communities.

Interviews with IFRC staff examined RCCE implementation, mechanisms of impact, as well as how activities operated in a specific context. Topic guides for health workers explored vaccination program delivery, influences on people’s decisions to take vaccines, (including SLL), and the extent to which community perspectives were incorporated. Topic guides for community members and leaders examined experiences with outbreaks, COVID-19, vaccination and the health system. Community participants were not necessarily familiar with the IFRC activities.

Citizen ethnography, a participatory method in which, typically lay community members ^30^, are trained to use ethnographic methods was used to generate locally grounded evidence. ^30^. RC volunteers received training in both ethnographic methods and positionality, given their prior roles in RCCE implementation. Observations were recorded using structured templates and discussed in debrief sessions with the research team.

Audio recordings were transcribed and translated into English. A coding tree was collaboratively developed between local and international research partners and IFRC/RC teams, and analysis was aided by NVivo software. A validation and cross-site analysis workshop was held in Abuja with all partners to refine the initial analysis and support interpretation of findings.

### Ethics

Formal ethics approval for this study was granted from the London School of Hygiene and Tropical Medicine Ethics Committee (29697), the University of Geneva Ethics Committee (CUREG-20230814-224-2) and the national Ethics Boards in Ethiopia (reference EPHA/OG/1078/24) and Nigeria (NHREC/01/01/2007-30/10/2023).

Written consent was given by all participants (or in the case of illiterate participants an independently witnessed thumb print) after being provided with study information in their own language and the opportunity to ask questions.

### Patient and public involvement

Participatory methods enabled participants, including vaccine users, to reflect on their own experiences of vaccination and community engagement. Red Cross volunteers were trained as citizen ethnographers and contributed to data collection and analysis. Early findings were shared and validated with participants and local stakeholders, including national Red Cross societies and partners. Research results were disseminated, and feedback sought, through local dissemination workshops in both Kano and Dire Dawa that included local health authorities, RC staff and volunteers and community association representative.

We used the SRQR reporting checklist^31^ when editing this manuscript, included in supplement 1.

## Findings

### Theory of Change for the RCCE intervention

In our research, we asked programme staff how they understood the SLL RCCE programme operating, for whom, under what circumstances and why.

The ‘theory of change’ for the existing RCCE intervention is depicted in Figure 2. According to participants, the main focus of RCCE activities was about sharing information about the vaccines **[‘activities’ in blue]** through various channels. These were ‘two-way’ channels such as house-to-house visits as well as ‘mass’ media communication. There was also an emphasis on community participation through consulting community leaders, for example about concerns around the vaccines. This was anticipated to strengthen knowledge of vaccines **[‘outputs’ in green]** and address concerns and counter misinformation on vaccines. The main mechanisms were knowledge based. Outputs were expected to be achieved because vaccine users were willing to act on their knowledge **[‘mechanisms’ in pink]**. This was also triggered by the existing trust of the institution, the IFRC/RC National Societies, and relational trust with individual RC volunteers and opinion leaders selected to promote the vaccine and share information. Feedback was expected to lead to adaptations to the IFRC’s vaccination messages. This would then contribute *to* improved uptake and coverage of the vaccines in Dire Dawa and Kano **[far right]**. The intervention operated in, and was influenced by a particular context **[‘context’ in red]**.i

**Figure 2:**
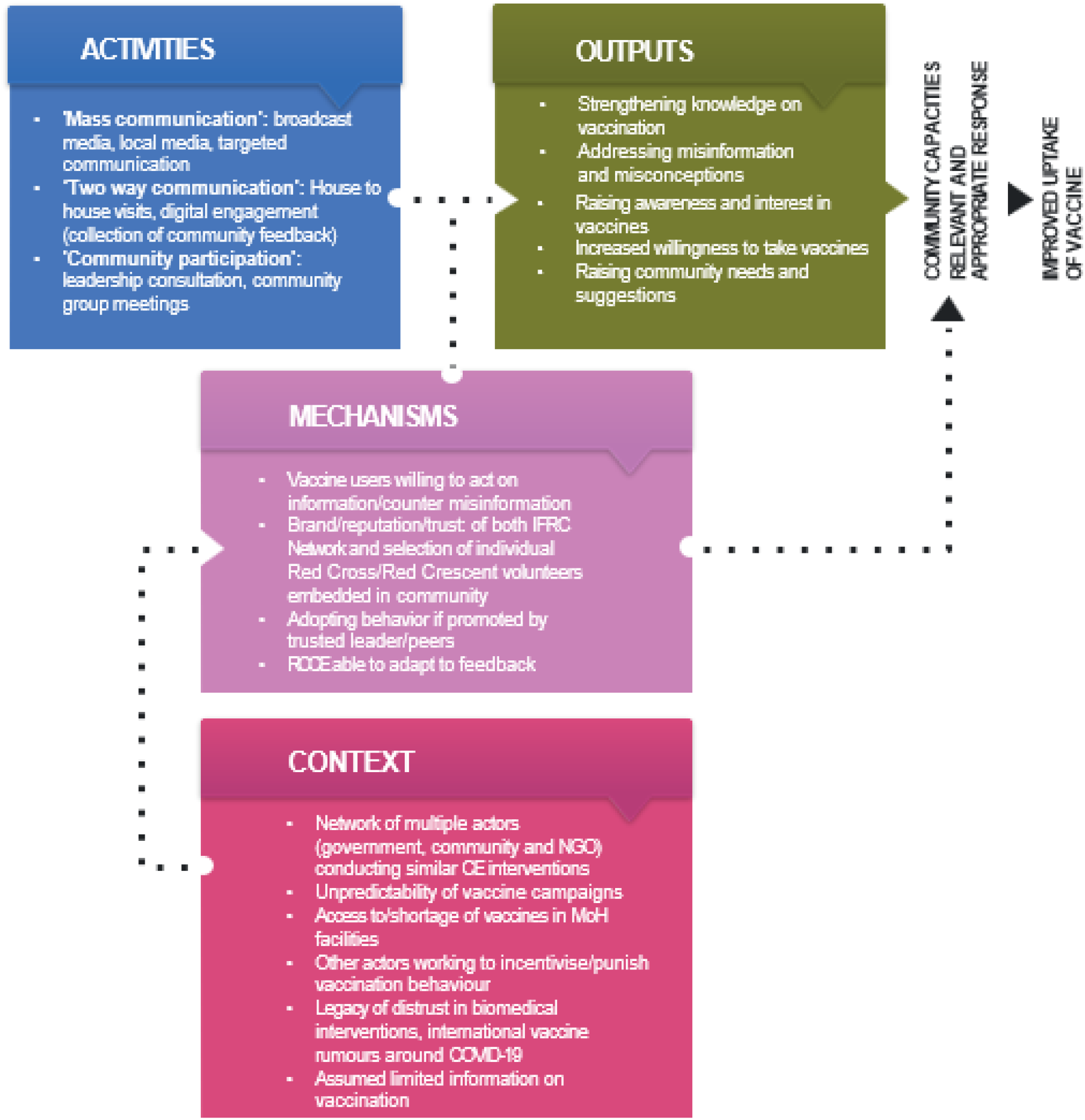
‘Theory of change’ for the IFRC Network’s RCCE intervention

## Understood mechanisms of change for RCCE

### Vaccine users willing to act on (mis)information

RCCE implementors often stated that their main goals were to tackle misinformation and debunk rumours. Respondents discussed COVID-19 rumours at length, including notions that vaccines cause infertility and are tools of population control, the vaccine containing micro-chips or concerns around rapid vaccine development. RCCE often addressed this with one way mass communication delivering correct biomedical information on vaccines. RC volunteers equipped with information on the safety, efficacy and benefits of taking a vaccine disseminated this information through the radio and town crier announcements. Providing this information was intended to increase uptake by improving people’s knowledge.

> *“The community is informed through various methods, such as loudspeakers placed in the town, announcements made by cars driving around the village, and different media outlets. However, there is a challenge because some people are resistant to accepting the information”* (IFRC Network staff, Ethiopia)

House-to-house visits by RC volunteers involved discussions (typically limited to a few minutes) with household heads about vaccination. These conversations often involved a two-way dialogue, as shown in this example of a member of staff trying to convince a community member that a rumour was false:

> *“Her children told her that if you receive COVID-19 vaccination and put a spoon on your arm then the spoon will stick to your body so therefore she will not accept vaccination…*.*then I said to her let us do a little, experiment…I placed the spoon on her arm and it stuck. I asked her again “did you receive COVID-19 vaccinations?” She said “no”, then I asked “why did it get stuck?”…I said “it is because your body is sticky because of the sweat”* (IFRC Network staff, Nigeria)

There were some examples where community feedback shaped the design or adaptation of vaccination delivery:

*“So for instance, if the feedback for hesitancy or not being vaccinated was the distance to the next vaccination point, then that helped to structure and say, ‘Oh, maybe we need mobile vaccine points that are closer to this communities’ because their main issue is not on the content of the vaccine per se, but it’s mostly more a question of accessing the site*.*”* (IFRC Network staff, Ethiopia)

However, this was unusual, and often communities participated at the point of implementation rather than being involved early on in the design of a vaccination strategy

### Adopting behaviour if promoted by trusted leader/peer

We found that trust was an important mechanism of change. The use of power brokers as well as influential or credible community members emerged as an important way through which the RCCE intervention may have led to outcomes. These leaders were identified and engaged by IFRC Network staff, provided with vaccine-related messages, and then supported to disseminate these messages through group sessions:

> *“You need to have people who are listened to… For example we worked with the higher official of industrial park, who has to be senior for his juniors…Then next we invited the religious leader since the community listen to the religious institution from Islam, Christian, Evangelical… We called them to deal with situation even there were even scientifically religious misconception. It is better to be clear about 666 [a number representing the devil] from the religious people than us* (IFRC Network staff, Ethiopia)

Our power mapping indicated that individuals or groups influencing vaccination can vary over time and differ by locality, this level of nuance may be difficult for a rapid vaccination campaign to capture.

### Implementers’ reputation/trust

The reputation of individual volunteers was important. Teams selected trusted individuals in the community who were locally embedded, such as nurses or educated individuals:

> *“Our community volunteers are community-based people, they are not visitors…he knows an outsider, he knows a bad person, he knows a good person, so he knows how to relate with them very well…they know when and where to enter and they know when and where to bring out results. That has been our strength.”*(IFRC Network staff, Nigeria)

Similarly, implementers mentioned the institutional reputation or ‘brand’ of the Red Cross/Red Crescent, particularly it’s neutrality, which may contrast with other authorities in countries marked by recent conflicts.

### Challenges and gaps identified

Limited community participation in the design of vaccination strategies created distinct challenges, including poor timing of campaigns:

> *We just sit down from here and say that we are going to do a measles campaign in the middle of September or August when people are in the farm, or we want to do a measles campaign when the schools are not in session or when the schools are in session*. (IFRC Network staff, Nigeria)

Staff also noted that a strong focus on information delivery limited meaningful community involvement in decision-making:

> *“The major problem is that we think the community isn’t aware, but the community is aware of their problems…*.*we don’t allow community members to participate*.
>
> *Sometimes we prepare the plan without the involvement of community members. But community members know more about their problem…Our role should be facilitation*..*” (IFRC Network staff, Ethiopia)*
>
> Tensions also arose where vaccination was a lower priority or competed with other needs in humanitarian settings:
>
> *“sometimes it will be difficult to address all community problems. For example, they say our problem is food, not COVID…COVID is not a community concern*.*” (IFRC Network staff, Ethiopia)*

## Context in which RCCE was taking place

### Crowded CE space

We identified a network of actors working on vaccine delivery and community engagement in both settings. Community members themselves led activities mirroring IFRC approaches, such as mass communication of vaccination messages, yet the SLL intervention was rarely integrated with these efforts. A community leader described some of these activities:

> *“once we get the information from the government, we disseminate the information to the ward heads to pass the message to their people, we also employ town announcers to go around the villages in the night to pass these messages, we also use the mosque to pass the messages to the people during prayer sessions*.*”* (Community leader, Nigeria)

### Health system and supply issues

Health system and supply challenges with vaccines were a defining feature of both contexts. For example, one facility in Dire Dawa lacked measles stock for one month. Equally common were reports of health workers refusing to open measles vaccine vials due to concerns about wastage, while caretakers described feeling fatigued from repeated attempts to access to vaccines.

> *“There could be no cold chain to save the vaccine…*.*you spend a lot of money on RCCE and it’s not even the RCCE that is the problem, the problem is that the vaccines are not even available*.*” (IFRC Network staff, Nigeria)*

### Unpredictability of government vaccine campaigns

RCCE took place alongside government campaigns with rapidly changing schedules. During our study period (January to May 2024), the IFRC Network often only had a few days’ notice to deliver an RCCE intervention before vaccine supplies arrived and were deployed, limiting opportunities for meaningful community involvement in planning and decision-making.

### Coercion and incentives

Participants also described coercive strategies, including enforcement by police and religious police authorities amongst households perceived as ‘resistant’ who rejected vaccination. This could take punitive forms, or in some cases, the costs involved in accessing vaccines were covered:

> *“So, once I rain down instructions on the respective ward head where we have cases of hesitancy, I will command the hesitant person to be summoned. So, others follow suit. But there are others who actually are in poverty and not that they don’t want to accept it [the vaccine], but they don’t have the means, such people with genuine cases are assisted by us*.*”*(Community leader, Nigeria)

Whilst this was potentially counter to global standard around community engagement, this appeared to be a localised way of implementing RCCE activities. In the case of Nigeria, there were also incentives reported for vaccination. An INGO-run conditional cash transfer programme operated whereby caretakers were paid around 1 USD upon presenting their infant for routine immunisation.

### Perceptions of vaccines

Aside rumours and concerns specific to COVID-19, there were also locally specific drivers of mistrust in vaccines. For example, Kano was the location of the 1996 Pfizer meningitis drug trial, which resulted in high profile lawsuits over the company’s failure to obtain informed consent from participants (Lenzer, 2006). Memories of this scandal were still mentioned in our study.

## Discussion

Despite renewed attention of its importance for humanitarian programming, significant gaps remain in understanding how community engagement works, for whom and in which contexts. By articulating a Theory of Change, we have shown how IFRC Network staff understood their RCCE intervention to work and the mechanisms through which they wanted to increase routine and emergency vaccination in Nigeria and Ethiopia. Our findings make two key contributions. Firstly, we interrogate this theory of change in relation to wider evidence and global narratives on vaccine confidence and the expected impact of community engagement. Teasing out existing gaps we propose that more thought needs to be given in the sector to explicitly articulating the mechanisms of change for community engagement and exploring how these can translate to changes in implementation. Secondly, taking the theory of change developed with our participants, we show that information delivery remains the predominant focus for RCCE practitioners.

In our research, RCCE implementers commonly aimed to combat misinformation and rumours, assuming this would increase vaccine coverage. This was highlighted by participants as a significant ‘mechanism’ in the theory of change we developed. This reflects global attention to the COVID-19 ‘infodemic’ and an emphasis on RCCE to ‘debunk’ rumours about safety and efficacy ^32^. However, this approach has however been critiqued, low vaccine uptake is rarely driven simply by poor information, but rather by mistrust in authorities and experts^32–35^. Assuming that community behaviour is to blame for low vaccination rates can distract from significant supply and health system access challenges that many people face, as our findings have shown.

Moving beyond ‘knowledge deficit’ approaches requires dialogical rather than didactic communication and mechanisms of change that focus more directly on building trust in institutions or implementers rather than debunking misinformation ^34,35^. Rather than seeing communities simply as passive recipients of vaccines, we need innovative approaches that envision community members actively co-creating vaccine delivery strategies with healthcare professionals ^36^. This means engaging communities to co-design interventions including identifying partners to extend reach, navigating security intelligence, and developing shared priorities that situate immunisation in communities’ broader concerns about health and wellbeing and overcoming uptake barriers. This is increasingly recognised by RCCE practitioners too, as two-way communication and even co-design are encouraged, however our findings show that translation into practice may be limited ^4^. For example, whilst we found some examples of community feedback being used to change elements of vaccination strategies this was challenging in an environment where government campaigns could be called at short notice, and where punitive approaches were taken with community members seen to resist vaccination.

Trust, in both the individual and institutional bearers of vaccine information, emerged as a key mechanism of change, This was based on the credibility, reputations and authority of implementors, volunteers conducting the RCCE, and the leaders or peers chosen to disseminate messages. We found that these ‘messengers’ may be more important than the messages themselves around vaccination. Whilst widely recognised as crucial for vaccine acceptance it is often not clear how humanitarian actors can facilitate trust. We found that influential leaders are context dependent and vary by locality, and RCCE teams responding to outbreaks may have very little time to investigate these dynamics. Power shifts over time and that its distribution is influenced by humanitarian interventions themselves, reinforcing the need for rapid and dynamic contextual analysis to accompany interventions whenever possible ^37,38^. Our study showed that qualitative methods such as power mapping can be rapidly deployed to identify trusted messengers^37,38^.

These insights raise questions about how to measure and understand outcomes beyond knowledge change. Future research and programme evaluation should articulate the diverse range of outcomes community engagement is trying to foster, this includes intermediate outcomes such as building trust in vaccine services, engagement of opinion leaders, or redistribution of decision making onto communities ^13^. Our research shows that engaging implementers and intended beneficiaries, can highlight tensions as well as opportunities in how community engagement is practiced, and can serve as a useful, empirical starting point from which to deliberate on how we can better align community engagement practice with existing evidence (e.g. on vaccine confidence) and normative goals such as co-production and dialogue.

### Strengths and limitations

Our findings are a result of collaborative research between Nigerian, Ethiopian and European research institutions and the IFRC and Red Cross/Red Crescent national societies. Regular meetings and workshops ensured that all partners contributed to the analysis and validation of this theory of change. This unique approach adds weight to the findings we have presented. However the study relied on retrospective accounts, and implementors may have overemphasized programme successes given their institutional roles.

## Conclusions

Our research with IFRC and the Red Cross/Red Crescent National Societies in Ethiopia and Nigeria contributes to the limited evidence base for community engagement practitioners seeking to design, evaluate or understand the impact of their programmes. While many RCCE interventions focus on communicating vaccine information and debunking misinformation, our study shows that trust in ‘messengers’ often plays a more critical role than the messages themselves. Future community engagement research and practice should focus on building a shared understanding of what the theory of change behind community engagement interventions ought to be. If community co-design, empowerment and ownership are expected to sit at the centre of community engagement practice, rather than delivering vaccine information to communities, a theory of change needs to reflect this and chart pathways to achieving this.

## Contributors

**RB:** conceptualisation, investigation, formal analysis, methodology, project administration, writing-original draft, writing-review and editing**; YH:** conceptualisation, investigation, formal analysis, methodology, data curation, project administration, writing-review and editing, supervision**; AD:** data curation, investigation, project administration, formal analysis**; YM:** investigation, project administration, formal analysis**; OB:** conceptualisation, formal analysis, writing-review and editing**; GF:** conceptualisation, formal analysis, writing-review and editing**; MP:** conceptualisation, formal analysis, writing-review and editing**; LA:** project administration, formal analysis**; AT:** project administration, formal analysis**; AO:** investigation, project administration, formal analysis**; KB:** conceptualisation, formal analysis, project administration, writing-review and editing, funding acquisition, supervision; **LE:** conceptualisation, formal analysis, project administration, writing original draft, writing-review and editing, funding acquisition, supervision.

## Funding

Our research was funded by the UK Humanitarian Innovation Hub. LE would also like to acknowledge support from the UK Research and Innovation (MR/T040521/1).

## Competing interests

The authors declare no conflicting interests in relation to this work.

## Patient consent for publication

All participants provided written informed consent for participation and for the anonymised use of their qualitative data in academic publications.

## Data availability statement

Given the sensitive and contextual nature of the qualitative data, we do not have permission from participants to share transcripts, however metadata and all other project documents are being deposited in the UK Data Service Archive.

## Acknowledgements

We would like to acknowledge the Red Cross staff and volunteers of Nigeria and Ethiopia Red Cross Society for their support and engagement throughout the study and especially those who participated as citizen ethnographers in Kano and Dire Dawa. We would also like to thank Rachel Cassidy, Wakgari Deressa, Abiy Seifu, Nanati Legese, Umar Tarajim, Sani Njobdi, Amina Nur Alkali for their support to data collection.

## Footnotes

1 Information on the Saving Lives and Livelihoods programmes can be found at: https://africacdc.org/saving-lives-and-livelihoods/

i For coherence within the IFRC Network, we have used the same terminology as the IFRC CEA impact framework (refer to Figure 1).

## Notes

### Competing Interest Statement

The authors have declared no competing interest.

